# Impact of different diagnostic measures on drug class association with dementia progression risk: a longitudinal prospective cohort study

**DOI:** 10.1101/2021.08.03.21261570

**Authors:** Daman Kaur, Magda Bucholc, David P. Finn, Stephen Todd, KongFatt Wong-Lin, Paula L. McClean

## Abstract

**Background:** Clinical Dementia Rating Sum of Boxes (CDRSOB) scale is known to be highly indicative of cognitive-functional status, but it is unclear whether it is consistent with clinical diagnosis in evaluating drug class associations with risk of progression to mild cognitive impairment (MCI) and dementia.

**Methods:** We employed multivariable logistic regression on longitudinal NACC data, to identify drug classes associated with disease progression risk, using clinical diagnosis and CDRSOB as the outcome.

**Results:** Anticoagulants, non-steroidal anti-inflammatory drugs, antihypertensives, antidepressants, and Parkinson’s medications were significantly associated with decreased progression to mild cognitive impairment (MCI)/dementia, and antipsychotics, antidiabetics, hypolipidemic drugs, and Alzheimer’s disease (AD) medications were significantly associated with increased progression risk. Associations were however dependant on the diagnostic measure used, e.g., levodopa was associated with reduced MCI-to-AD risk using CDRSOB as the outcome (OR:0.28, FDR p<0.002), but not with clinical diagnosis. Additionally, some associations appear to be gender specific; for instance, antiadrenergic agents had lower MCI-to-Dementia risk only for men (OR:0.67, FDR p<0.001) using CDRSOB.

**Conclusions:** Overall, we demonstrate that choice of diagnostic measure can influence the magnitude and significance of risk or protection attributed to drug classes. A consensus must be reached within the research community with respect to the most accurate diagnostic outcome to identify risk and improve reproducibility.

## Introduction

Dementia is a complex disease with several subtypes and aetiologies, and few effective therapeutics (Arvanitakis et al., 2019). Therefore, extensive research has been carried out analysing factors that may influence disease incidence and progression (Livingston et al., 2020; Peters et al., 2019). Medications, in particular, may affect cognition in older adults (Barton et al., 2008; Nikaido et al., 1990) due to increased drug sensitivity associated with age-related factors such as impaired liver metabolism and decreased renal function (Barton et al., 2008). Moreover, factors like potentially inappropriate prescribing, drug interactions and polypharmacy, which are common in dementia patients, complicate the assessment of specific drug classes on cognition (Delgado et al., 2019).

Several pharmacoepidemiology studies have analysed the relationship between medications and dementia risk with conflicting findings. Researchers have acknowledged that methodological differences substantially contribute to the variation in risk attributed to different medications. Study design, inclusion criteria, data preparation, and especially the diagnostic criteria used can influence outcomes (Erkinjuntti et al., 1997; Jamsen et al., 2016; Wancata et al., 2007). One study examining antidiabetic medications found protective effects of metformin on dementia risk (Chin-Hsiao 2019), whereas another reported increased risk of cognitive impairment associated with metformin use (Moore et al., 2013). For antihypertensives, one systematic review reported a significant association between reduced dementia risk and use of diuretics and angiotensin-converting enzyme (ACE) inhibitors (Shah et al., 2009), whereas a study by Rouch et al. (2015) showed that calcium (Ca^2+^) channel blockers and renin–angiotensin system blockers, were associated with prevention of dementia. Similarly, discrepant findings across studies have been reported on the association between anxiolytics, such as benzodiazepines, and cognitive decline (Salzman, 2020; Verdoux et al., 2005). This is also the case for other drug classes including antidepressants, antipsychotics, hypolipidemic drugs, and non-steroidal anti-inflammatory (NSAIDs) (Biringer et al., 2009; Hill et al., 2010; Imbimbo et al., 2010; Schulz et al., 2018; Zhang et al., 2018). We note significant variability in the diagnostic criteria used across studies that undoubtedly impacts upon risk attribution leading to conflicting findings.

Clinical assessment of dementia involves detailed examination of medical history, cognitive tests followed by laboratory assays, psychiatric evaluation, and brain imaging to identify dementia subtype. Studies have shown that subtle differences in classification associated with National Institute of Neurological and Communicative Disorders and Stroke-Alzheimer’s Disease and Related Disorders Association (NINCDS-ADRDA), Diagnostic and Statistical Manual Of Mental Disorders (DSM) criteria including DSM–III–R, DSM–IV, DSM-V, International Classification of Diseases (ICD) criteria including ICD-9, ICD-10, ICD-11, and Cambridge Mental Disorders of the Elderly Examination (CAMDEX) criteria can influence clinical diagnosis, and lead to variation in dementia prevalence (Berman & Bursztajn 1999; Chaves et al., 2007; Erkinjuntti et al., 1997; Wancata et al., 2007). As a result of this variation, researchers often employ cognitive scores for analysis, especially in longitudinal studies as they allow for identifying small changes in cognition over time. However, in clinical settings cognitive tests are mainly used to aid the diagnostic decision-making process.

The Clinical Dementia Rating (CDR^®^ Dementia Staging Instrument) scale, a cognitive assessment tool, is regularly used in clinical and research settings to gauge dementia severity. It provides a global score, and a more detailed CDR sum of boxes (CDRSOB) score obtained through patient and informant interview based on the following cognitive and functional domains; memory, orientation, judgment & problem solving, community affairs, home & hobbies, and personal care (Morris, 1993). Studies have reported moderate (Forsell et al., 1992; Juva et al., 1994), to good (Chaves et al., 2007; Ding et al., 2018; Lima et al., 2017) correlation of the CDR scale with DSM and McKhann (1984) diagnostic criteria, with one study reporting efficiency of CDRSOB in distinguishing MCI from dementia for patients with CDR global score 0.5 (O’Bryant et al.,2010). However, it is unclear whether such cognitive-functional assessments are consistent with clinical diagnosis in terms of evaluating the benefits or risks of medications.

In this study, we investigated how the associations between medications and dementia risk vary between clinical diagnosis and CDRSOB scores generally, and differentially in men and women (Trenaman et al., 2009). We analysed several drug classes available in the National Alzheimer’s Coordinating Center (NACC) dataset, namely, antihypertensives, lipid lowering medication, NSAIDs, anticoagulants, antidepressants, antipsychotics, anxiolytics, antidiabetics, Alzheimer’s disease (AD) medication and Parkinson’s disease (PD) medication, and subcategories of drugs within each class, for identifying those significantly associated with progression to MCI, all-cause dementia, and AD.

## Methods

### Data source

Archival data from the National Alzheimer’s Coordinating Center (NACC), consisting of over 500 variables on genetic, lifestyle, and clinical features for 39,531 individuals was used in this study. Details about the NACC, recruitment of participants, and assessment process has been previously described (Besser et al., 2018; Morris et al., 2006).

The NACC Uniform Data Set (UDS), comprising of data collected from September 2005 until September 2020 was used in our analysis. The NACC is approved by the University of Washington Institutional Review Board. Written, informed consent from all participants and co-participants included in NACC-UDS was obtained by the Alzheimer’s disease research centers (ADRCs) where they completed their study visits. For the purpose of publication patient consent was not required. The following drug classes were analysed: antihypertensives, lipid lowering medication, NSAIDs, anticoagulants/antiplatelets, antidepressants, antipsychotic agents, anxiolytics/sedatives/hypnotics, diabetes medication, AD medication, and Parkinson’s disease medication. The diagnostic category of participants was determined based on both clinical diagnosis and CDRSOB scores. In NACC-UDS Version 1 and 2, the process of clinical diagnosis for all-cause dementia relied on the diagnostic protocol of the ADRC, with centres generally using DSM-IV (1994) or NINCDS-ADRDA guidelines. In NACC-UDS Version 3, the criteria for all-cause dementia were modified from McKhann (2011). Diagnoses of MCI were established using the modified Petersen criteria. Individuals included in the analyses were aged ≥40 years.

|  | **Diagnostic criteria for**<br>**UDS version 1.0 – 2.0** | **Diagnostic criteria for**<br>**UDS version 3.0** |
| --- | --- | --- |
| **Mild cognitive impairment (MCI)** | Petersen & Morris, 2005 | Petersen & Morris, 2005 |
| **Alzheimer's disease (AD)** | NINCDS-ADRDA criteria (McKhann et al., 1984) | NIA-AA criteria (McKhann et al., 2011) |
| **All-cause dementia** | Not specified (McKhann et al., 1984 or DSM-IV) | McKhann et al., 2011 |

### Data preparation

A multi-time-point data preparation approach was employed to conduct progression analysis for reducing bias associated with variables measured at a single time point. The NACC dataset comprises routinely collected data which is updated roughly every three months and the time between visits for individuals ranges from 6 months to ∼5 years. However, the majority of participants are followed up every 1-2 years. Clinical diagnosis and CDRSOB scores were identified across all visits for each participant to determine changes in cognitive status over time. Subsequently, six groups were identified, and the following five comparisons were assessed for both diagnostic measures; participants that were healthy across all visits (Remained Healthy), participants that were MCI across all visits (Remained MCI), those who progressed from Healthy-to-MCI, from Healthy-to-AD, from MCI-to-Dementia, and from MCI-to-AD. Individuals with single visits or a fluctuating diagnosis were excluded, and only those with a clear progression trend were analysed. CDRSOB scores were categorized as healthy: 0, MCI: 0.5-4.0 and dementia: 4.5-18 (O’Bryant et al., 2010).

Since covariates like diabetes, traumatic brain injury can occur after baseline, observations across all visits before the individual progressed to MCI/dementia were examined to determine their presence/absence. In the case of education and number of years smoked, the maximum value across all visits before the individual progressed to MCI/dementia was used for analysis. Body mass index (BMI) was categorized as underweight-1 (<18.5 kg/m2), normal-2 (18.5–24.99 kg/m2), or overweight-3 (>24.99 kg/m2). Subsequently, transitions in BMI over time were determined by calculating the average of BMI categories (underweight, normal, or overweight) across all visits and qualitatively comparing this to the baseline category to determine increase/decrease/stable progression (Kaur et al., 2020). Figure 1 depicts the sample selection process for the different progression groups analysed. Details about the multi-time-point analysis approach have been published previously (Kaur et al., 2020).

**Fig. 1:**
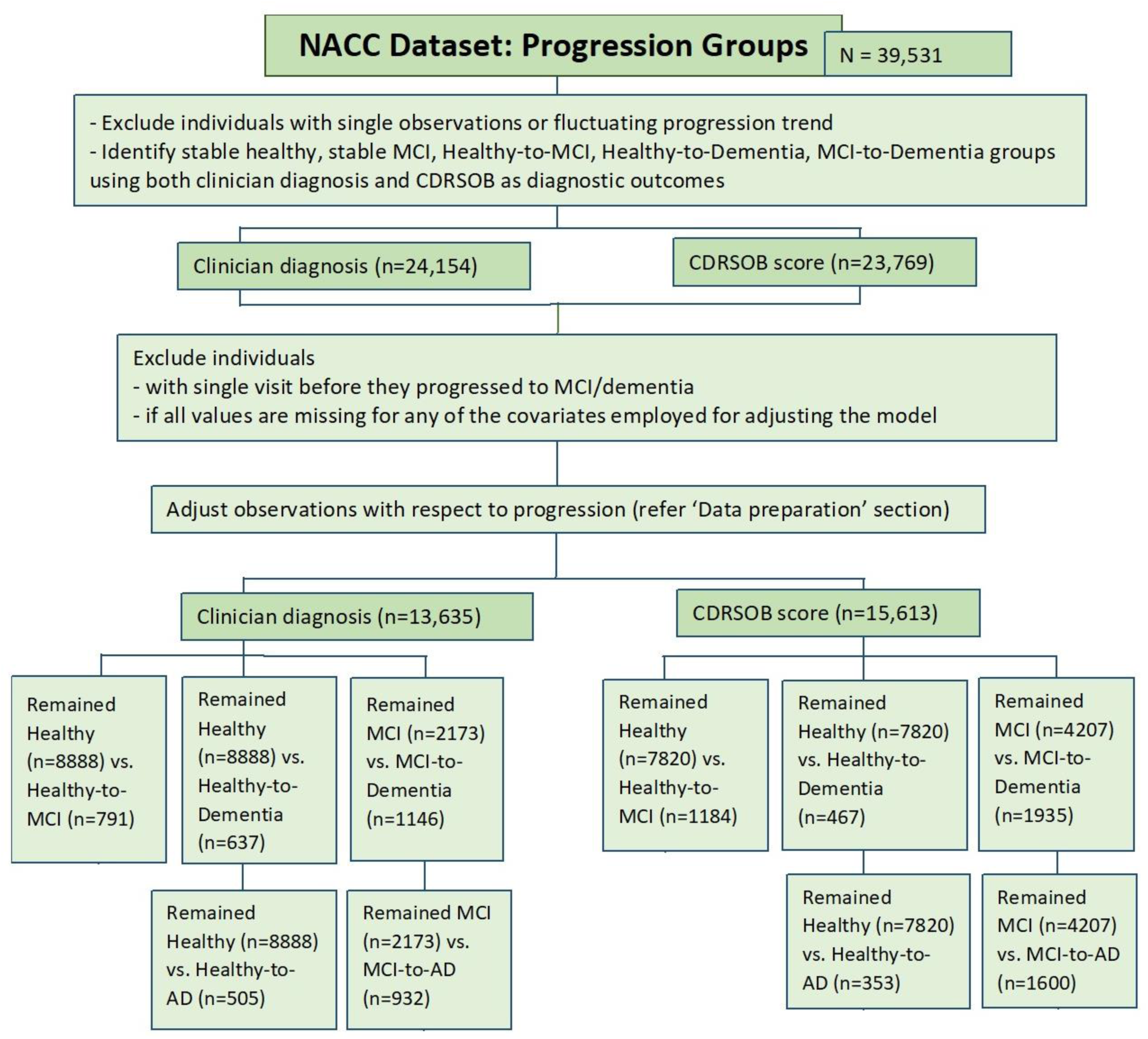
Data preparation process for progression groups within the NACC dataset. (AD: Alzheimer’s Disease; CDRSOB: Clinical Dementia Rating Sum of Boxes; MCI: Mild Cognitive Impairment; NACC: National Alzheimer’s Coordinating Center).

The major drug classes analysed in this study, and subcategories of antihypertensives are available in the UDS dataset as variables. For subcategorising the other significant drug classes, we used the researcher’s data dictionary (RDD; Kukull, 2015) to identify the common drug names stored in the UDS. Next, we searched for specific drug terms (Supplementary File 1) across all visits to determine prescription data for each individual, those who reported a specific drug at least during one visit were classified as taking the medication. These were then categorised based on their mechanism of action. Medications with ophthalmic and topical routes of administration were excluded from this analysis. Table 1 represents the drug classes and their respective subcategories that were analysed in this study. Surprisingly, it was observed that some cognitively healthy individuals (n=70) were taking AD medication, either a reflection of genuine prescribing patterns for AD drugs or errors at the point of data entry. These data were removed and not analysed.

**Table 1:**
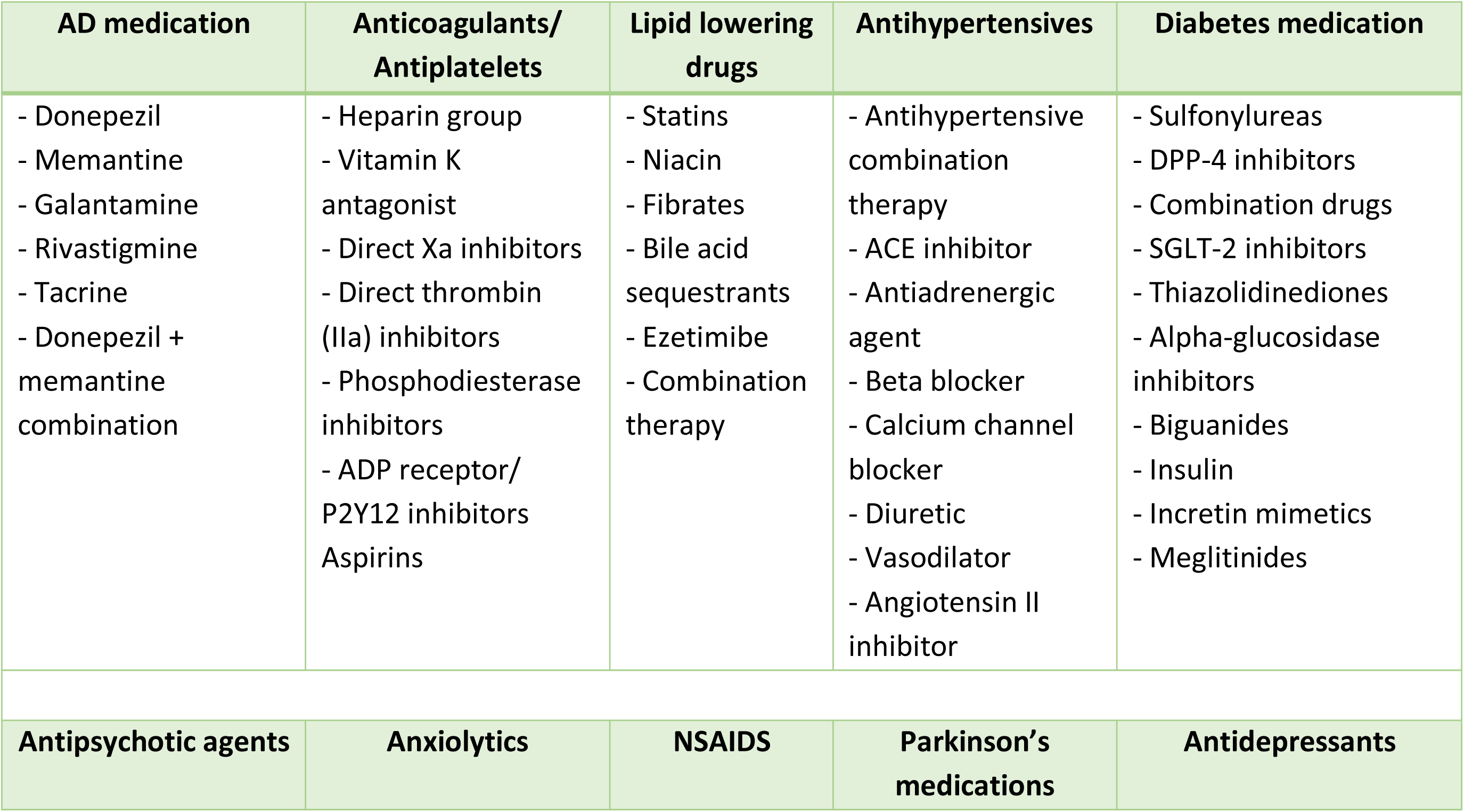
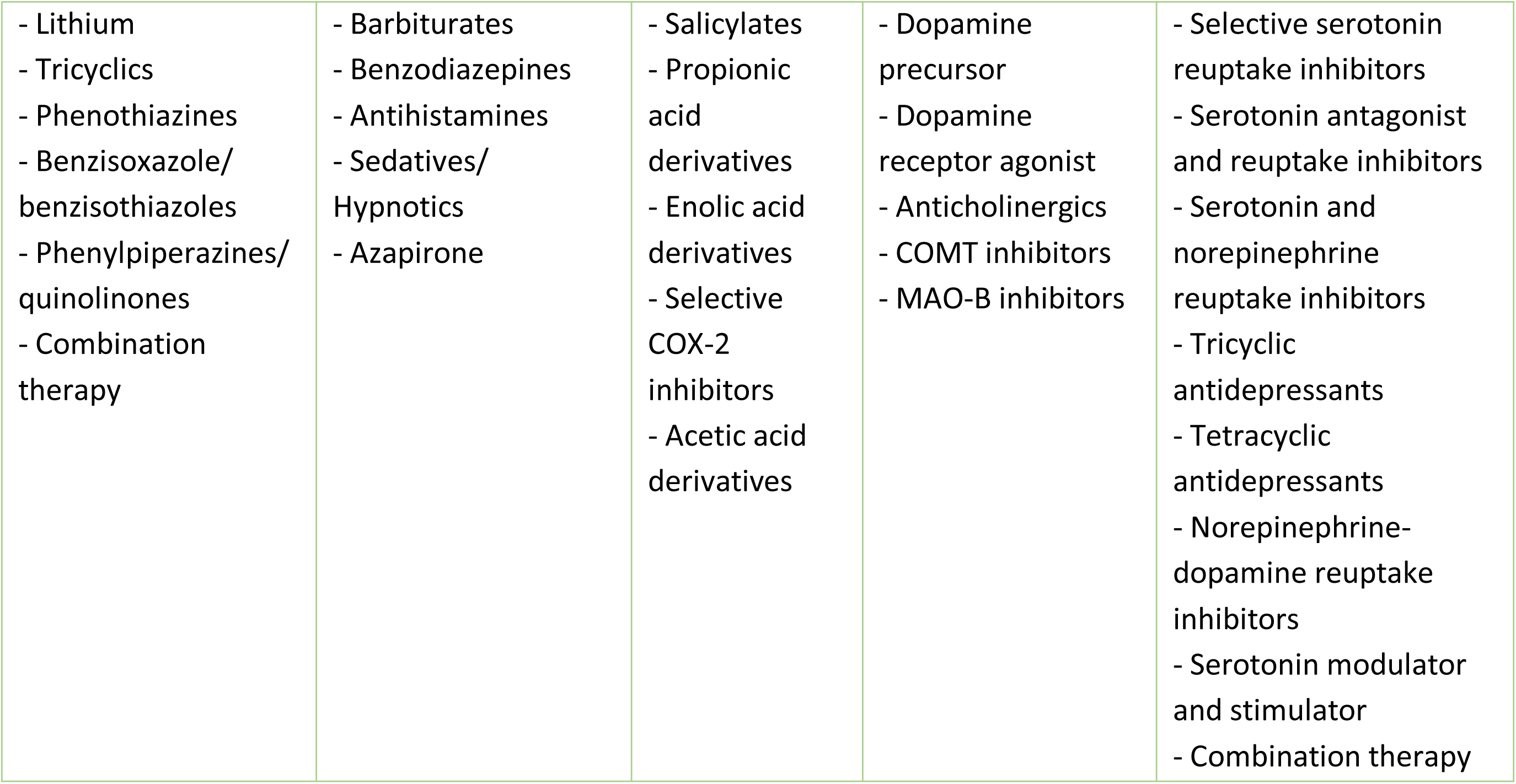
Drug classes analysed, and their subcategories based on the mechanism of action. (AD: Alzheimer’s Disease; ADP: Adenosine Diphosphate; ACE: Angiotensin-converting enzyme; DPP-4: Dipeptidyl peptidase-4; SGLT-2: Sodium-glucose co-transporter-2; NSAIDs: Nonsteroidal anti-inflammatory drugs; COX-2: Cyclooxygenase-2; COMT: Catechol-O-methyltransferase; MAO-B: Monoamine oxidase type B).

| AD medication | Anticoagulants/<br>Antiplatelets | Lipid lowering<br>drugs | Antihypertensives | Diabetes medication |
| --- | --- | --- | --- | --- |
| - Donepezil<br>- Memantine<br>- Galantamine<br>- Rivastigmine<br>- Tacrine<br>- Donepezil + memantine combination | - Heparin group<br>- Vitamin K antagonist<br>- Direct Xa inhibitors<br>- Direct thrombin (IIa) inhibitors<br>- Phosphodiesterase inhibitors<br>- ADP receptor/P2Y12 inhibitors<br>- Aspirins | - Statins<br>- Niacin<br>- Fibrates<br>- Bile acid sequestrants<br>- Ezetimibe<br>- Combination therapy | - Antihypertensive combination therapy<br>- ACE inhibitor<br>- Antiadrenergic agent<br>- Beta blocker<br>- Calcium channel blocker<br>- Diuretic<br>- Vasodilator<br>- Angiotensin II inhibitor | - Sulfonylureas<br>- DPP-4 inhibitors<br>- Combination drugs<br>- SGLT-2 inhibitors<br>- Thiazolidinediones<br>- Alpha-glucosidase inhibitors<br>- Biguanides<br>- Insulin<br>- Incretin mimetics<br>- Meglitinides |
| Antipsychotic agents | Anxiolytics | NSAIDS | Parkinson's medications | Antidepressants |
| - Lithium<br>- Tricyclics<br>- Phenothiazines<br>- Benzisoxazole/benzisothiazoles<br>- Phenylpiperazines/quinolinones<br>- Combination therapy | - Barbiturates<br>- Benzodiazepines<br>- Antihistamines<br>- Sedatives/Hypnotics<br>- Azapirone | - Salicylates<br>- Propionic acid derivatives<br>- Enolic acid derivatives<br>- Selective COX-2 inhibitors<br>- Acetic acid derivatives | - Dopamine precursor<br>- Dopamine receptor agonist<br>- Anticholinergics<br>- COMT inhibitors<br>- MAO-B inhibitors | - Selective serotonin reuptake inhibitors<br>- Serotonin antagonist and reuptake inhibitors<br>- Serotonin and norepinephrine reuptake inhibitors<br>- Tricyclic antidepressants<br>- Tetracyclic antidepressants<br>- Norepinephrine-dopamine reuptake inhibitors<br>- Serotonin modulator and stimulator<br>- Combination therapy |

### Statistical analysis

Shapiro-Wilk test was employed to assess normality of data for demographic analysis. In the case of continuous variables, significance of differences was evaluated using an independent t-test for normally distributed data, and the Mann-Whitney U test for non-normally distributed data. For categorical variables, a chi-square test was applied compare differences.

In order to explore the associations between different drug classes and disease progression, multivariable logistic regression was employed (Glonek et al., 1995). Major drug classes were first analysed together, followed by analysis of their subcategories (Table 1). Additionally, the analysis was repeated for men and women separately, to determine sex-specific effects associated with drug exposure. The analyses were adjusted for important confounders and risk factors of dementia including age, sex, race, years of education and smoking, alcoholism, hypertension, diabetes, cardiovascular disorders, depression, BMI, traumatic brain injury, and hearing impairment (Livingston et al., 2020). Additionally, to determine whether significant associations occur due to the medication or the underlying condition, respective comorbidities were also included in the analysis, for e.g., Parkinson’s disease (PD) for analysing PD medications, schizophrenia and bipolar disorder for analysing antipsychotics.

Sensitivity analysis was performed by analysing individuals who reported a specific drug at least during two visits. Adjustment for multiple hypothesis testing was achieved by applying false discovery rate (FDR) using the Benjamini-Yekutieli correction method (Benjamini & Yekutieli, 2001). FDR adjusted p-values (FDR p) <0.01 were considered statistically significant.

### Software and codes

Statistical analyses were performed using the ‘PredictABEL’ package in R studio (Version 1.1.423) on a Windows machine with eight memory cores. Codes are available on GitHub (web link to be made publicly available upon acceptance of manuscript).

## Results

### General characteristics

The average follow-up time for the groups analysed using clinical diagnosis (CDRSOB) was 4.92 (4.86) years for stable healthy, 5.82 (5.65) years for Healthy-to-MCI, 2.65 (2.94) years for stable MCI, 6.4 (6.88) years for Healthy-to-Dementia, and 3.65 (3.8) years for MCI-to-Dementia progression group.

Analysis revealed that individuals who progressed from Healthy-to-MCI and from Healthy-to-Dementia were less likely to be married (p=0.01, p=0.01, respectively), a higher proportion of them suffered from conditions like hypertension (p<0.0001, p<0.0001, respectively), cardiovascular disorders (p<0.0001, p<0.0001, respectively), depression (p<0.05, p<0.0001, respectively), and hearing impairment (p<0.0001, p<0.0001, respectively), and they had a higher average of total years smoked compared to stable healthy individuals (p<0.05, p=0.001, respectively). Those who progressed from Healthy-to-Dementia were also significantly less educated (p<0.0001), and a higher proportion of them reported decrease in BMI over time compared to stable healthy individuals (p<0.0001).

Compared to the stable MCI group, those who progressed from MCI-to-Dementia were more likely to be married (p<0.0001), a higher proportion of them suffered from depression (p<0.05) and hearing impairment (p<0.05), and a lower proportion suffered from diabetes (p<0.001) and hypertension (p<0.0001). Detailed results can be found in Supplementary File 2.

### Medication analysis

Using multivariable logistic regression, we identified drug classes associated with risk of progression from Healthy-to-MCI, Healthy-to-Dementia, MCI-to-Dementia, and MCI-to-AD for both CDRSOB and clinical diagnosis as the outcome. The adjusted odds ratios were calculated based on drug exposure vs. absence of the drug of interest during the timeline available within the NACC dataset. For brevity, significant results are illustrated in Figures 2-5. Complete results, including the insignificant associations, can be found in Supplementary File 3.

**Fig. 2:**
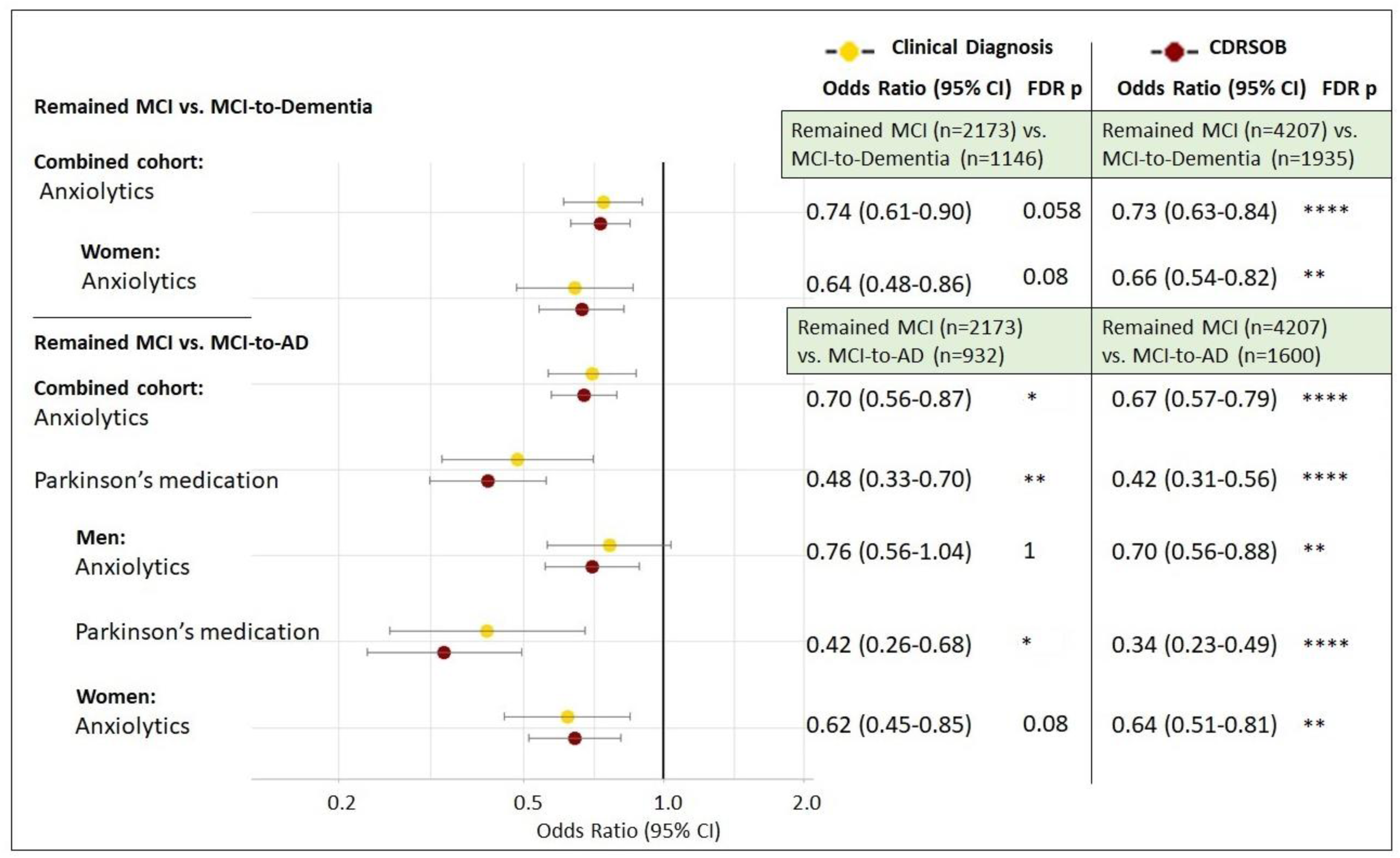
Summary grouped forest plots of adjusted odds ratios for drug classes significantly associated with risk of progression to dementia and AD for two diagnostic variables; clinical diagnosis and CDRSOB scores. (AD: Alzheimer’s Disease; MCI: Mild Cognitive Impairment; CDRSOB: Clinical Dementia Rating sum of boxes; CI: Confidence Intervals). *\*p<0*.*05, **p<0*.*01, ***p<0*.*001, ****p<0*.*0001*.

### Parkinson’s medications are associated with reduced MCI-to-AD progression risk in men

Comparing stable MCI individuals to those who progressed from MCI-to-Dementia/AD, anxiolytics were significantly associated with decreased progression risk overall, and in men and women separately only with CDRSOB scores as the outcome (FDR p<0.01; Fig. 2). Parkinson’s medications on the other hand, were associated with reduced MCI-to-AD risk overall (FDR p<0.01), and specifically for men (FDR p<0.01) with both clinical diagnosis and CDRSOB scores as the outcome (Fig. 2). None of the major drug classes analysed were significantly associated with risk of progression from Healthy-to-MCI or Healthy-to-Dementia/AD.

### Antiadrenergic agents are associated with reduced MCI to dementia progression risk in men

We then focused on analysis of subcategories of the drug families. In terms of anticoagulants, direct Xa inhibitors were significantly associated with reduced Healthy-to-MCI risk (FDR p=0.001) overall with CDRSOB as the outcome (Fig. 3A). They were also associated with reduced MCI-to-Dementia and MCI-to-AD risk with CDRSOB as the outcome, however with a lower significance level (FDR p<0.05; Fig. 3A). Dopamine precursors that are used to treat Parkinson’s disease, were significantly associated with reduced MCI-to-AD risk overall (FDR p<0.01) and specifically in men with CDRSOB as the outcome (FDR p<0.05; Fig. 3A). Furthermore, antiadrenergic agents, a type of antihypertensive medication, were associated with reduced MCI-to-Dementia and MCI-to-AD risk overall (FDR p<0.001), and specifically in men with CDRSOB as the outcome (FDR p<0.01; Fig. 3B).

**Fig. 3:**
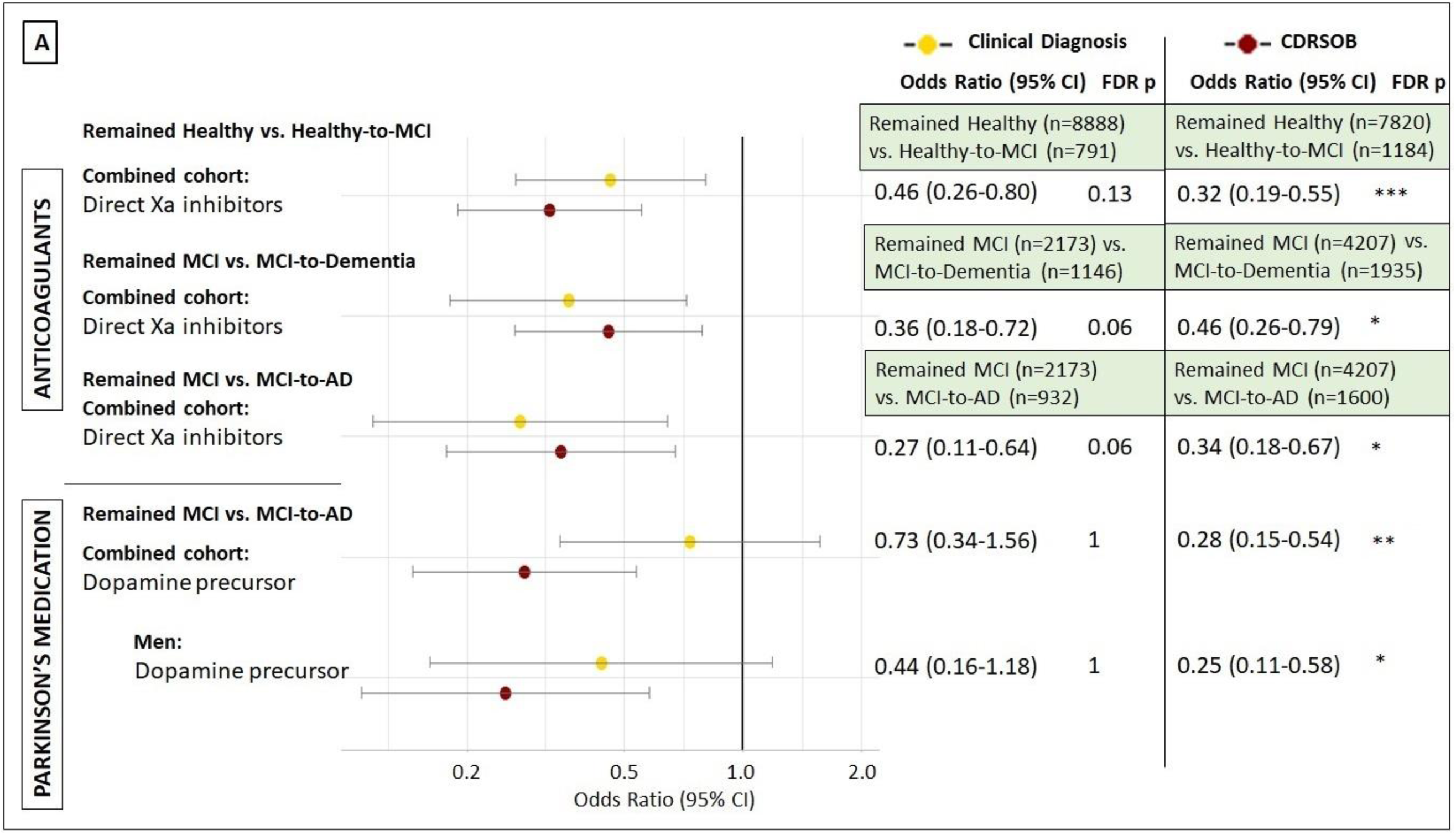
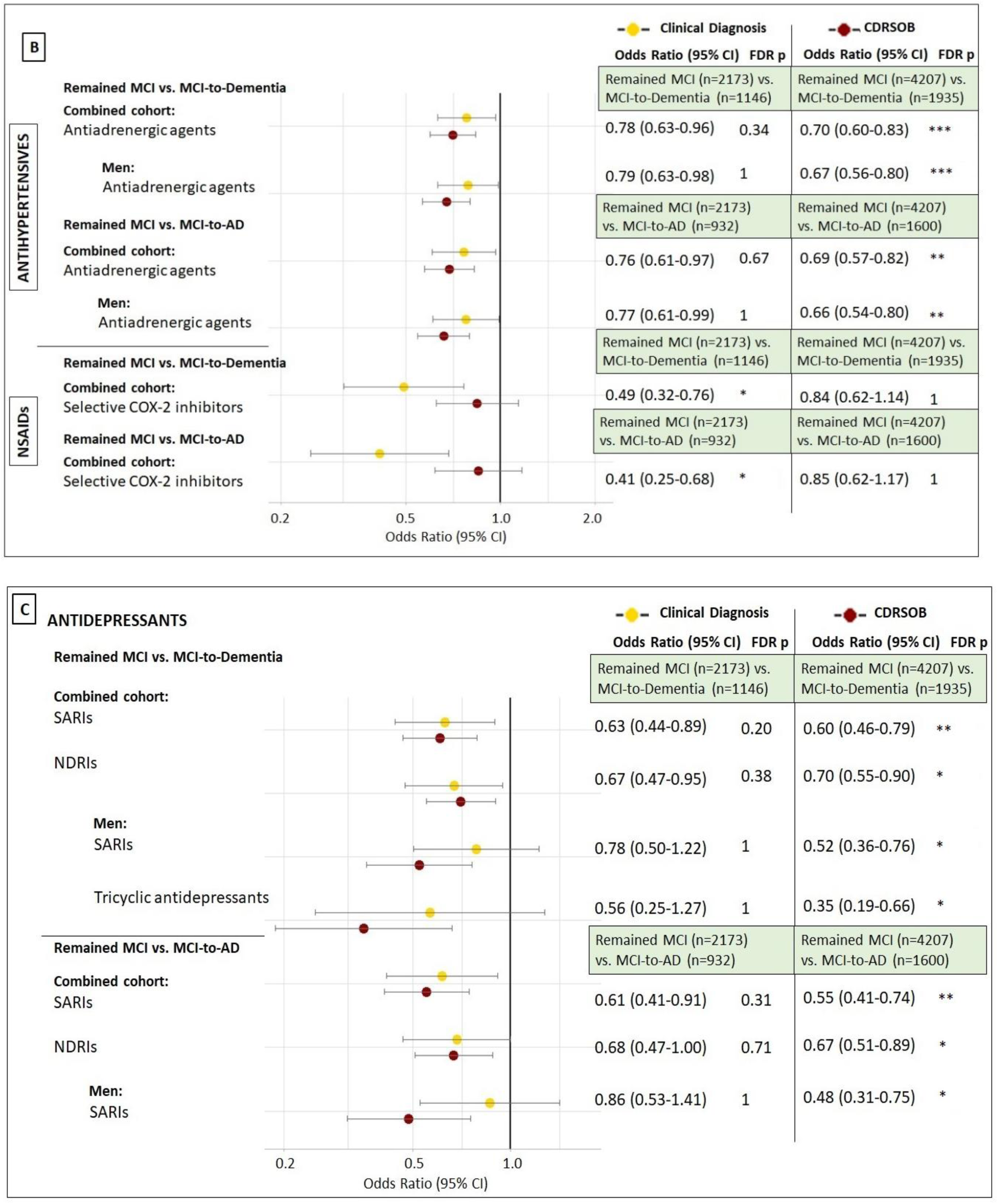
Summary grouped forest plots of adjusted odds ratios for drug subcategories significantly associated with reduced risk of progression to MCI, dementia, and AD for two diagnostic variables; clinical diagnosis and CDRSOB scores. Drug classes analysed: (A) Anticoagulants and Parkinson’s medication; (B) Antihypertensives and NSAIDs; (C) Antidepressants. (AD: Alzheimer’s Disease; CDRSOB: Clinical Dementia Rating sum of boxes; CI: Confidence Intervals; COX: Cyclooxygenase-2; MCI: Mild Cognitive Impairment; NDRIs: Norepinephrine-dopamine reuptake inhibitors; SARIs: Serotonin antagonist and reuptake inhibitors). *\*p<0*.*05, **p<0*.*01, ***p<0*.*001, ****p<0*.*0001*.

While analysing subcategories of NSAIDs, selective COX-2 inhibitors were associated with reduced MCI-to-Dementia and MCI-to-AD risk overall with clinical diagnosis as the outcome (FDR p<0.05; Fig. 3B). In terms of antidepressants, Serotonin antagonist and reuptake inhibitors (SARIs; FDR p<0.01) and Norepinephrine-dopamine reuptake inhibitors (NDRIs; FDR p<0.05) were significantly associated with reduced MCI-to-Dementia and MCI-to-AD progression risk overall with CDRSOB as the outcome (Fig. 3C). Further, SARIs and tricyclic antidepressants were associated with reduced MCI-to-Dementia risk in men with CDRSOB as the diagnostic outcome (FDR p<0.05; Fig. 3C).

Cardiovascular disorders, hypertension, and depression are known to be associated with impaired cognition. The difference in significance observed between CDRSOB and clinical diagnosis for these drug classes may represent improved cognition associated with drug treatment, however these medications may not play a role in reducing the risk of dementia. In the case of dopamine precursors, co-occurrence of AD and PD is infrequent, however similar cognitive-functional domains are affected in both disorders which might explain the significant results obtained with CDRSOB scores (Fig. 3B).

### Combination hypolipidemic therapy and thiazolidinediones are associated with increased MCI-to-AD risk with CDRSOB as outcome

Analysis of lipid lowering drugs revealed that combination therapy was associated with increased MCI-to-Dementia and MCI-to-AD risk with CDRSOB as the outcome (FDR p<0.05; Fig. 4). Additionally, in the case of antidiabetic medication, thiazolidinediones were associated with increased MCI-to-AD risk with CDRSOB as the diagnostic measure (FDR p<0.05; Fig. 4).

**Fig. 4:**
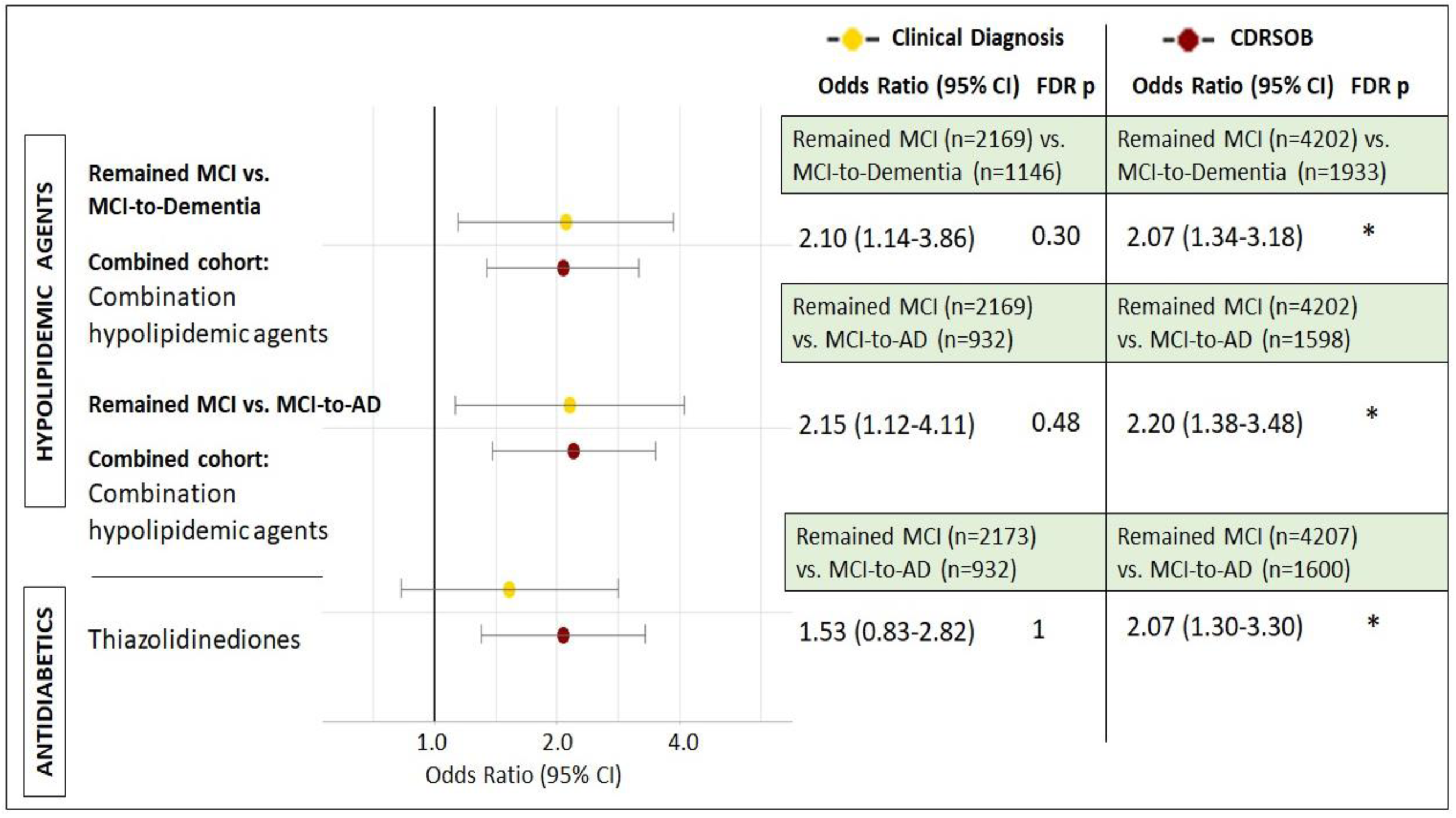
Summary grouped forest plots of adjusted odds ratios for drug subcategories significantly associated with increased risk of progression to dementia and AD for two diagnostic variables; clinical diagnosis and CDRSOB scores. Drug classes analysed: Hypolipidemic agents and Diabetes medication. (AD: Alzheimer’s Disease; CDRSOB: Clinical Dementia Rating sum of boxes; CI: Confidence Intervals; MCI: Mild Cognitive Impairment). *\*p<0*.*05*.

### AD drug treatments in individuals with MCI are associated with increased risk of dementia and AD

Commonly prescribed AD medications were analysed in participants who remained MCI over time vs. those who progressed from MCI to AD specifically or dementia in general. Donepezil, a cholinesterase inhibitor (ChEI) was significantly associated with increased MCI-to-Dementia risk (FDR p<0.001; Fig. 5A) and MCI-to-AD risk (FDR p<0.001; Fig. 5B) in general, and in men and women with both clinical diagnosis and CDRSOB as the outcome. Similarly, memantine, an N-methyl-D-aspartate (NMDA) receptor antagonist was also significantly associated with increased MCI-to-Dementia and MCI-to-AD risk overall and for men and women with both clinical diagnosis and CDRSOB (FDR p<0.001; Fig. 5).

**Fig. 5:**
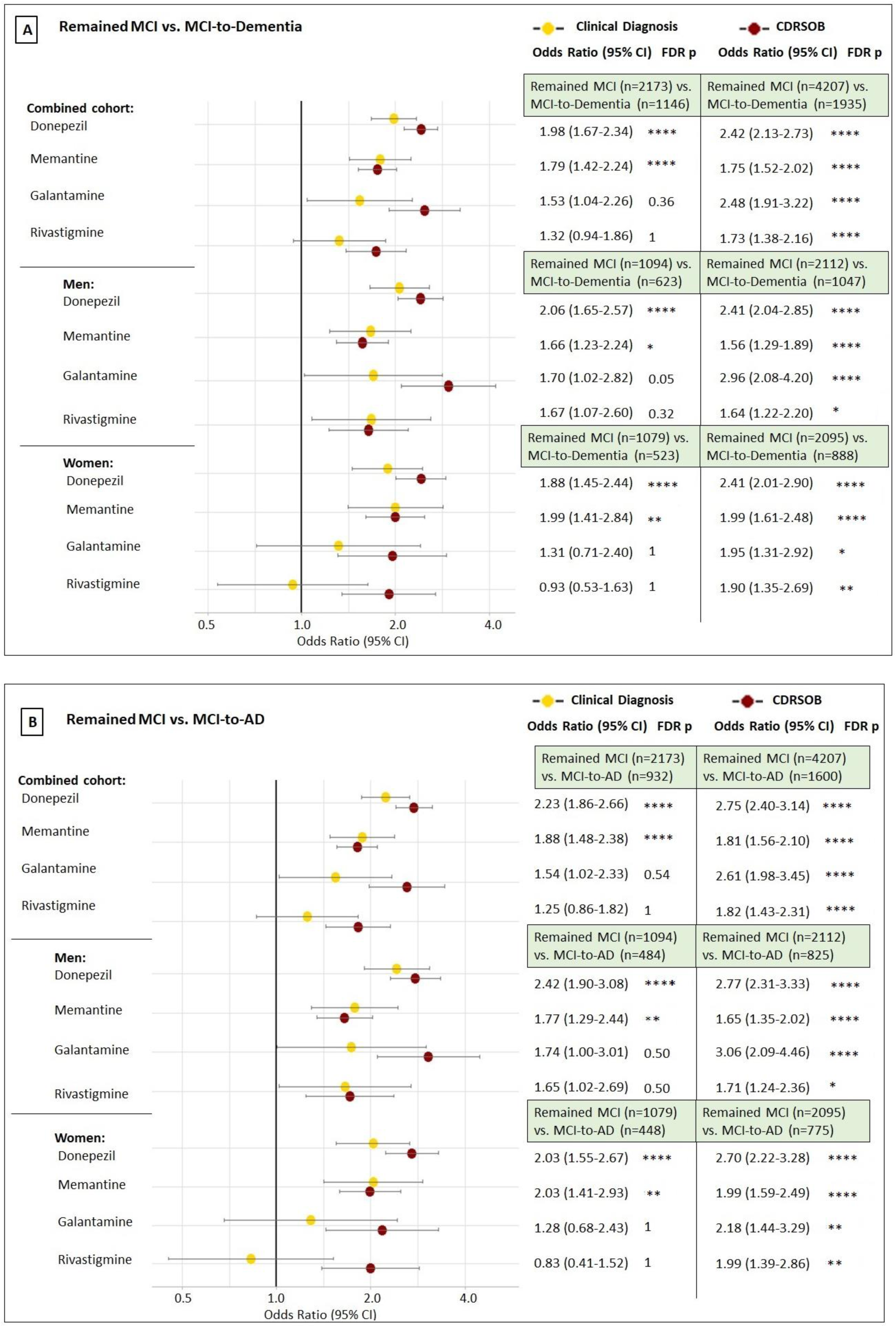
Summary grouped forest plots of adjusted odds ratios for AD drugs significantly associated with risk of progression to dementia and AD for two diagnostic variables; clinical diagnosis and CDRSOB scores. Progression groups analysed: (A) Remained MCI vs. MCI-to-Dementia; (B) Remained MCI vs. MCI-to-AD. (AD: Alzheimer’s Disease; CDRSOB: Clinical Dementia Rating sum of boxes; CI: Confidence Intervals; MCI: Mild Cognitive Impairment). *\*p<0*.*05, **p<0*.*01, ***p<0*.*001, ****p<0*.*0001*.

Galantamine and Rivastigmine (both ChEIs), were associated with increased MCI-to-Dementia and MCI-to-AD progression risk in general (FDR p<0.001), and for both men and women with CDRSOB as the diagnostic measure, but not clinical diagnosis (Fig. 5).

In the NACC dataset, more individuals are classified as MCI or dementia with CDRSOB scores as compared to clinical diagnosis (Fig. 5). Further, higher proportion of individuals are prescribed donepezil/memantine compared to galantamine/rivastigmine which may explain the difference in significance of results for the latter while comparing the two diagnostic outcomes.

### Sensitivity Analysis

A more conservative approach was adapted for performing sensitivity analysis by only including individuals who reported a specific medication during at least two visits. The results obtained from sensitivity analysis were in accordance with the primary analysis. Anxiolytics (OR: 0.58, FDR p<0.01) and PD medications (OR: 0.26, FDR p<0.001) were significantly associated with reduced MCI-to-AD risk. Direct Xa inhibitors were associated with reduced Healthy-to-MCI risk (OR: 0.58, FDR p<0.01). Antiadrenergics (OR: 0.71, FDR p=0.04) and SARIs (OR: 0.52, FDR p<0.01) were associated with reduced MCI-to-Dementia risk. Further, dopamine precursors were associated with reduced MCI-to-AD risk (OR: 0.09, FDR p=0.03). Sensitivity analysis of AD medications also revealed significant results that were in accordance with the primary analysis for both CDRSOB and clinical diagnosis (Supplementary File 3).

In the case of selective COX-2 inhibitors, NDRIs, tricyclic antidepressants, hypolipidemic combination therapy, and thiazolidinediones which had FDR p<0.05 in the primary analysis (Figures 3-4), results from sensitivity analysis, although not statistically significant were qualitatively similar to the primary analysis. The detailed regression results can be found in Supplementary File 3.

## Discussion

In this study, we demonstrate that the use of different diagnostic measures for MCI and dementia, i.e., clinical diagnosis and CDRSOB scores, affect the significance of the association of various medications with disease risk and progression. From the drug classes analysed, anxiolytics and Parkinson’s Disease (PD) medications were found to be differentially associated with risk of progression to all-cause dementia and AD with respect to the diagnostic measure used. Additionally, subcategories of drug classes including direct Xa inhibitors, dopamine precursors, antiadrenergic agents, selective COX-2 inhibitors, SARIs, NDRIs, and tricyclic antidepressants were associated with decreased progression risk, whereas combination hypolipidemic therapy and thiazolidinediones were associated with increased progression risk.

In the NACC dataset, consensus-based clinical diagnosis was made using the criteria proposed by McKhann et al. (Mckhann et al., 1984; Mckhann et al., 2011). A previous study, analysing the NACC dataset, reported diagnostic accuracy with 0.71 sensitivity and 0.81 specificity when comparing CDRSOB scores to dementia diagnosis (O’Bryant et al., 2010). We report that despite good correlation between the two diagnostic measures (Ding et al., 2018), our analysis produced discordant results while analysing risk associated with different medications. Whilst odds ratios were consistently in the same direction, the statistical significance of results was inconsistent for the two diagnostic measures.

The CDRSOB scale offers a wider score range allowing better demarcation of subtle changes between different stages of disease progression. In the NACC cohort, a lower number of individuals are classified into Healthy-to-MCI group by clinical diagnosis (n=791), compared to CDRSOB score (n=1184; Fig. 1). This suggests that CDRSOB scale can detect early disease symptoms and help diagnose MCI. Whereas in terms of a clinical diagnosis, there are no specific tests that confirm MCI, judgement is made based on clinical evaluation and exclusion of other causative factors such as hypothyroidism or vitamin B12 deficiency. These factors not only affect the prevalence of MCI and dementia in a cohort but may also lead to variation in risk analysis.

In our study, PD medications were significantly associated with reduced MCI-to-AD risk overall, and specifically for men given that PD is more common in males, with CDRSOB as the outcome (FDR p<0.0001). Dopamine precursors, in particular, were associated with significantly reduced MCI-to-AD risk with CDRSOB as the outcome. Studies have shown that levodopa treatment can have beneficial effect on cognition (Ikeda et al., 2017). Additionally, PD related MCI is clinically distinct and has a longer conversion period to dementia, compared to AD related MCI (Besser et al., 2016). In the case of anxiolytics, while they were significantly associated with reduced MCI-to-AD progression risk (Fig. 2), analysis of its subclasses did not reveal any significant results. However, anxiety itself was associated with reduced MCI-to-AD risk (OR: 0.54, FDR p<0.05; Supplementary File 3).

Direct factor Xa inhibitors, which are prescribed for treatment and prevention of blood clots and strokes were significantly associated with reduced Healthy-to-MCI and MCI-to-Dementia/AD risk. These results are in accordance with several published studies that found reduced risk of cognitive impairment and dementia associated with oral anticoagulants (Cheng et al., 2018; Ding et al., 2018; Zeng et al., 2019). Direct Xa inhibitors in particular are associated with lower dementia risk compared to other anticoagulants (Lee et al., 2021).

Antiadrenergic agents were significantly associated with reduced MCI-to-Dementia/AD risk overall, and specifically for men with CDRSOB scores (FDR p<0.01). Several studies have reported increased risk of cognitive impairment associated with hypertension, and a subsequent reduced risk with antihypertensive treatment (Ding et al., 2020; Shah et al., 2009). Although there is ambiguity regarding the duration, specific drug subclass, and optimal dosage to prevent cognitive impairment (Ou et al., 2020). A previous study analysing the NACC dataset reported lower cognitive scores associated with lower systolic BP (Fiford et al., 2020). Analysis of antidepressants revealed significant association of SARIs with reduced MCI-to-Dementia/AD progression risk (FDR p<0.01). While depression had an OR value of 1.32 (FDR p<0.001) suggesting that it is associated with increased risk of progression, however, treatment of the disorder with certain antidepressants can protect against this risk. Studies have suggested that SARIs like trazodone exhibit neuroprotective properties when examined in mammalian cell cultures and in neurodegenerative mouse models (Halliday et al., 2017) with complex cellular signalling pathway mechanisms (Joshi et al., 2020).

With respect to NSAIDs, conflicting outcomes have been reported in observational and experimental studies (Ali et al., 2019; Breitner et al., 2009; Cote et al., 2012). Our analysis showed reduced MCI-to-Dementia/AD risk associated with selective COX-2 inhibitors while using clinical diagnosis. These results are in accordance with preclinical studies that found beneficial effects of selective COX-2 inhibitors on memory and synaptic plasticity, as it has been shown to hinder the neurotoxic effects of amyloid beta (Kotilinek et al., 2008; Woodling et al., 2016). NSAID use may also lead to modulation of inflammatory pathways which can be beneficial as inflammatory genes and molecules have been linked with AD development (Newcombe et al., 2018; Raj et al., 2017). Additionally, studies have shown that some NSAIDs can mediate Aβ clearance. However, it is only effective during the early disease stages (Imbimbo et al., 2010).

We found that thiazolidinediones were associated with increased MCI-to-AD risk with CDRSOB as the outcome (FDR p<0.05). Despite the association of diabetes with lower cognitive function, several studies have reported a reduced dementia risk in those taking oral hypoglycaemic medication (Hsu et al., 2011; Kim et al., 2019; Wu et al., 2020). However, in the case of thiazolidinediones, drug exposure has been associated with faster cognitive decline and poorer immediate memory (Seaquist et al., 2013; Wu et al., 2020). Increased risk of dementia linked to intensive diabetes treatment leading to hypoglycaemia has also been reported (Zammitt & Frier, 2005). In terms of lipid lowering drugs, statin combination therapy was also associated with increased MCI-to-Dementia progression risk with CDRSOB (FDR p<0.05).

While focusing on specific AD drugs for MCI-to-Dementia and MCI-to-AD progression, donepezil and memantine were significantly associated with increased risk of disease progression with both clinical diagnosis and CDRSOB as the outcome. However, galantamine and rivastigmine were found to be significantly associated only with CDRSOB as the outcome. Previous studies have shown faster cognitive decline with ChEI therapy in MCI and early AD (Han et al., 2019; Schneider et al., 2011). These associations could be due to confounding by indication since patients with cognitive impairment are more likely to be prescribed these drugs. It also reflects the prescribing patterns across clinics since, in contradiction to the US FDA guidelines, physicians are known to prescribe ChEIs and memantine during early stages of impairment (Schneider et al., 2011).

Overall, our study shows that the magnitude and significance of association of different medications with disease progression can vary depending on the diagnostic measure used. CDRSOB scores, though comparatively less comprehensive, form a part of clinical diagnosis which leads to high correlation between the two. Due to unavailability of clinical diagnosis in some datasets, or a preference for more quantitative analysis to increase precision and reduce subjectivity, researchers often opt for cognitive scores (Bucholc et al., 2019; Ding et al., 2018; McCombe et al., 2022). However, we must be aware of the implications of choosing a particular diagnostic measure and be careful regarding how we interpret the results.

There were limitations in this study, such as the low number of individuals while analysing subcategories of drug classes. Importantly, it should be noted that significant results obtained in this study are associations and not causation, as the data was not generated from randomised controlled trials. Therefore, our findings are vulnerable to unmeasured confounders or confounding by indication, with AD drugs for example. Moreover, in this study exposure to the drug classes was analysed irrespective of duration, dosage, and drug-drug interactions. To address these issues there is a need for more detailed studies on larger cohorts with data on dosage, that will allow analysis of specific drug classes individually, while making use of comprehensive drug interaction databases like DDInter (Xiong et al., 2022). Undoubtedly, further studies are required to identify the most relevant diagnostic measure and to inform prescribing patterns in order to reduce modifiable risk of dementia. Additionally, based on our results we believe that further studies should focus on a gender stratified approach as there may be differential risk profiles associated with various subcategories of drug classes in men and women.

## Data Availability

The processed data can be found in the results section in the manuscript and Supplementary Table. R codes for data preparation and analysis will
be uploaded to GitHub in due course. Data source: NACC asks investigators to not share the data with individuals who are not collaborators on
the project for which the data was requested. This is partially due to the fact that they have distinctions between commercial and noncommercial recipients in place due to the option for NACC participants to elect to decline sharing of their data with commercial entities. Additionally, NACC has a data use agreement in place to help prevent misuse of the data. Lastly, this is helpful in the tracking of proposals, publications and data requests. NACC data is available through request to any interested researcher regardless of commerciality. At the time of the
request submission, investigators will be asked to provide details of the proposal and will need to submit a data use agreement. Requests can
be submitted on the website (https://naccdata.org/requesting-data/submit-data-request).

## Legends for Supplementary Files

<u>Supplementary File 1</u>: Medication terms used for identifying drug exposure across patient visits, split by drug classes and drug subcategories.

<u>Supplementary File 2</u>: Demographic characteristics of stable and progression groups analysed with clinician diagnosis and CDRSOB scores as diagnostic outcomes.

<u>Supplementary File 3:</u> Detailed regression results

Supplementary Table 3A: Detailed regression results for stable Healthy vs. Healthy-to-MCI analysis (*p<0.05, **p<0.01, ***p<0.001, ****p<0.0001).

Supplementary Table 3B: Detailed regression results for stable Healthy vs. Healthy-to-Dementia analysis (*p<0.05, **p<0.01, ***p<0.001, ****p<0.0001).

Supplementary Table 3C: Detailed regression results for stable Healthy vs. Healthy-to-AD analysis (*p<0.05, **p<0.01, ***p<0.001, ****p<0.0001).

Supplementary Table 3D: Detailed regression results for stable MCI vs. MCI-to-Dementia analysis (*p<0.05, **p<0.01, ***p<0.001, ****p<0.0001).

Supplementary Table 3E: Detailed regression results for stable Healthy vs. MCI vs. MCI-to-AD analysis (*p<0.05, **p<0.01, ***p<0.001, ****p<0.0001).

## Acknowledgements

1. This work was supported by the European Union’s INTERREG VA Programme, managed by the Special EU Programmes Body (SEUPB (Centre for Personalised Medicine, IVA 5036)), with additional support by the Northern Ireland Functional Brain Mapping Project Facility (1303/101154803), funded by invest Northern Ireland and the University of Ulster (K.W.-L.), Alzheimer’s Research UK (ARUK) NI Pump Priming (M.B.,S.T.,K.W.-L.,P.L.M.), Ulster University Research Challenge Fund (M.B.,S.T.,K.W.-L.,M.B.), and the Dr George Moore Endowment for Data Science at Ulster University (M.B.). The views and opinions expressed in this paper do not necessarily reflect those of the European Commission or the SEUPB.
2. The NACC database is funded by NIA/NIH Grant U01 AG016976. NACC data are contributed by the NIA-funded ADCs: P30 AG019610 (PI Eric Reiman, MD), P30 AG013846 (PI Neil Kowall, MD), P50 AG008702 (PI Scott Small, MD), P50 AG025688 (PI Allan Levey, MD, PhD), P50 AG047266 (PI Todd Golde, MD, PhD), P30 AG010133 (PI Andrew Saykin, PsyD), P50 AG005146 (PI Marilyn Albert, PhD), P50 AG005134 (PI Bradley Hyman, MD, PhD), P50 AG016574 (PI Ronald Petersen, MD, PhD), P50 AG005138 (PI Mary Sano, PhD), P30 AG008051 (PI Thomas Wisniewski, MD), P30 AG013854 (PI M. Marsel Mesulam, MD), P30 AG008017 (PI Jeffrey Kaye, MD), P30 AG010161 (PI David Bennett, MD), P50 AG047366 (PI Victor Henderson, MD, MS), P30 AG010129 (PI Charles DeCarli, MD), P50 AG016573 (PI Frank LaFerla, PhD), P50 AG005131 (PI James Brewer, MD, PhD), P50 AG023501 (PI Bruce Miller, MD), P30 AG035982 (PI Russell Swerdlow, MD), P30 AG028383 (PI Linda Van Eldik, PhD), P30AG053760 (PI Henry Paulson, MD, PhD), P30 AG010124 (PI John Trojanowski, MD, PhD), P50 AG005133 (PI Oscar Lopez, MD), P50 AG005142 (PI Helena Chui, MD), P30 AG012300 (PI Roger Rosenberg, MD), P30 AG049638 (PI Suzanne Craft, PhD), P50 AG005136 (PI Thomas Grabowski, MD), P50 AG033514 (PI Sanjay Asthana, MD, FRCP), P50 AG005681 (PI John Morris, MD), P50 AG047270 (PI Stephen Strittmatter, MD, PhD).”

## References

Ali, M.M., Ghouri, R.G., Ans, A.H., Akbar, A. & Toheed, A. 2019, “Recommendations for Antiinflammatory Treatments in Alzheimer’s Disease: A Comprehensive Review of the Literature”, Cureus, vol. 11, no. 5, pp. e4620.

American Psychiatric Association. 1994, “Diagnostic and statistical manual of mental disorders: DSM-IV” Washington, pp.866.

Arvanitakis, Z., Shah, R.C., Bennett, D.A. 2019, “Diagnosis and Management of Dementia: Review”, JAMA, vol. 322, no. 16, pp. 1589–1599.

Barton C., Sklenicka J., Sayegh P. & Yaffe, K. 2008, “Contraindicated medication use among patients in a memory disorders clinic.”, American Journal Geriatric Pharmacotherapy, vol. 6, no. 3, pp. 147–152.

Benjamini, Y. & Yekutieli, D. 2001, “The Control of the False Discovery Rate in Multiple Testing under Dependency”, The Annals of Statistics, vol. 29, no. 4, pp. 1165–1188.

Berman, S., Bursztajn, H.J. 1999, “Clinical criteria for three types of dementia had low sensitivity and high specificity.” Evidence-Based Mental Health, vol. 2, pp. 91.

Besser, L., Kukull, W., Knopman, D.S., Chui, H., Galasko, D., Weintraub, S., Jicha, G., Carlsson, C., Burns, J., Quinn, J., Sweet, R.A., Rascovsky, K., Teylan, M., Beekly, D., Thomas, G., Bollenbeck, M., Monsell, S., Mock, C., Zhou, X.H., Thomas, N., Robichaud, E., Dean, M., Hubbard, J., Jacka, M., Schwabe-Fry, K., Wu, J., Phelps, C., Morris, J.C. & Neuropsychology Work Group, Directors, and Clinical Core leaders of the National Institute on Aging-funded US Alzheimer’s Disease Centers 2018, “Version 3 of the National Alzheimer’s Coordinating Center’s Uniform Data Set”, Alzheimer Disease & Associated Disorders, vol. 32, no. 4, pp. 351–358.

Besser L.M., Litvan I., Monsell S.E., Mock C., Weintraub S., Zhou X.H. & Kukull, W. 2016, “Mild cognitive impairment in Parkinson’s disease versus Alzheimer’s disease.”, Parkinsonism and Related Disorders, vol. 27, pp. 54–60.

Biringer, E., Rongve, A. & Lund, A. 2009, “A Review of Modern Antidepressants’ Effects on Neurocognitive Function”, Current Psychiatry Reviews, vol. 5, no. 3, pp. 164–174.

Breitner, J.C.S., Haneuse, S.J.P.A., Walker, R., Dublin, S., Crane, P.K., Gray, S.L. & Larson, E.B. 2009, “Risk of dementia and AD with prior exposure to NSAIDs in an elderly community-based cohort SYMBOL SYMBOL”, Neurology, vol. 72, no. 22, pp. 1899–1905.

Bucholc, M., Ding, X., Wang, H., Glass, D. H., Wang, H., Prasad, G., Maguire, L. P., Bjourson, A. J., McClean, P. L., Todd, S., Finn, D. P., & Wong-Lin, K. (2019). A practical computerized decision support system for predicting the severity of Alzheimer’s disease of an individual. Expert systems with applications, 130, 157–171.

Cheng, W., Liu, W., Li, B., & Li, D. (201. Relationship of Anticoagulant Therapy With Cognitive Impairment Among Patients With Atrial Fibrillation: A Meta-Analysis and Systematic Review. Journal of cardiovascular pharmacology, 71(6), 380–387.

Chin-Hsiao, T. 2019, “Metformin and the Risk of Dementia in Type 2 Diabetes Patients.”, Aging & Disease, vol. 10, no. 1, pp. 37–48.

Chaves, M.L.F., Camozzato, A.L., Godinho, C., Kochhann, R., Schuh, A., de Almeida, V.L. & Kaye, J. 2007, “Validity of the clinical dementia rating scale for the detection and staging of dementia in Brazilian patients.”, Alzheimer Disease & Associated Disorders, vol. 21, no. 3, pp. 210–217.

Cote S., Carmichael P.H., Verreault R., Lindsay J., Lefebvre J. & Laurin, D. 2012, “Nonsteroidal anti-inflammatory drug use and the risk of cognitive impairment and Alzheimer’s disease.”, Alzheimer’s and Dementia, vol. 8, no. 3, pp. 219–226.

Delgado, J., Bowman, K. & Clare, L. 2020, “Potentially inappropriate prescribing in dementia: a state-of-the-art review since 2007”, BMJ Open, vol. 10, no. 1, pp. e029172.

Ding, J., Davis-Plourde, K. L., Sedaghat, S., Tully, P. J., Wang, W., Phillips, C., Pase, M. P., Himali, J. J., Gwen Windham, B., Griswold, M., Gottesman, R., Mosley, T. H., White, L., Guðnason, V., Debette, S., Beiser, A. S., Seshadri, S., Ikram, M. A., Meirelles, O., Tzourio, C., … Launer, L. J. (2020). Antihypertensive medications and risk for incident dementia and Alzheimer’s disease: a meta-analysis of individual participant data from prospective cohort studies. The Lancet. Neurology, 19(1), 61–70.

Ding, M., Fratiglioni, L., Johnell, K., Santoni, G., Fastbom, J., Ljungman, P., Marengoni, A., & Qiu, C. 2018. Atrial fibrillation, antithrombotic treatment, and cognitive aging: A population-based study. Neurology, 91(19), e1732–e1740.

Ding, X., Bucholc, M., Wang, H., Glass, D.H., Wang, H., Clarke, D.H., Bjourson, A.J., Dowey, L.R.C., O’Kane, M., Prasad, G., Maguire, L. & Wong-Lin, K. 2018, “A hybrid computational approach for efficient Alzheimer’s disease classification based on heterogeneous data.”, Scientific Reports, vol. 8, no. 1, pp. 9774.

Erkinjuntti, T., Ostbye, T., Steenhuis, R. & Hachinski, V. 1997, “The effect of different diagnostic criteria on the prevalence of dementia.”, New England Journal of Medicine, vol. 337, no. 23, pp. 1667–1674.

Fiford C.M., Nicholas J.M., Biessels G.J., Lane C.A., Cardoso M.J. & Barnes, J. 2020, “High blood pressure predicts hippocampal atrophy rate in cognitively impaired elders.”, Alzheimer’s and Dementia: Diagnosis, Assessment and Disease Monitoring, vol. 12, no. 1) (pagination, pp. Arte Number: e12035. ate of Pubaton: 2020.

Fink, H.A., Jutkowitz, E., McCarten, J.R., Hemmy, L.S., Butler, M., Davila, H.M.P.A., Ratner, E., Calvert, C., Barclay, T.R., Brasure, M.M.S.P.H., M.L.I.S., Nelson V.A. & Kane, R.L. 2018, “Pharmacologic Interventions to Prevent Cognitive Decline, Mild Cognitive Impairment, and Clinical Alzheimer-Type Dementia: A Systematic Review”, Annals of Internal Medicine, vol. 168, no. 1, pp. 39–51.

Gallacher, J., Elwood, P., Pickering, J., Bayer, A., Fish, M. & BenShlomo, Y. 2012, “Benzodiazepine use and risk of dementia: evidence from the Caerphilly Prospective Study (CaPS)”, Journal of Epidemiology & Community Health, vol. 66, no. 10, pp. 869–873.

Glonek, G.F.V., McCullagh, P. 1995, “Multivariate Logistic Models.” Journal of the Royal Statistical Society, vol. 57, pp. 533–546.

Halliday, M., Radford, H., Zents, K., Molloy, C., Moreno, J. A., Verity, N. C., Smith, E., Ortori, C. A., Barrett, D. A., Bushell, M., & Mallucci, G. R. 2017. Repurposed drugs targeting eIF2α-P-mediated translational repression prevent neurodegeneration in mice. Brain : a journal of neurology, 140(6), 1768–1783.

Han, J., Besser, L.M.M.S.P.H., Xiong, C., Kukull, W.A. & Morris, J.C. 2019, “Cholinesterase Inhibitors May Not Benefit Mild Cognitive Impairment and Mild Alzheimer Disease Dementia”, Alzheimer Disease & Associated Disorders, vol. 33, no. 2, pp. 87–94.

Hill, K.S., Bishop, J.R., Palumbo, D. & Sweeney, J.A. 2010, “Effect of second-generation antipsychotics on cognition: current issues and future challenges”, Expert Review of Neurotherapeutics, vol. 10, no. 1, pp. 43–57.

Hsu, C., Wahlqvist, M.L., Lee, M. & Tsai, H. 2011, “Incidence of dementia is increased in type 2 diabetes and reduced by the use of sulfonylureas and metformin.”, Journal of Alzheimer’s Disease, vol. 24, no. 3, pp. 485–493.

Ikeda M., Kataoka H. & Ueno, S. 2017, “Can levodopa prevent cognitive decline in patients with Parkinson’s disease?.”, American Journal of Neurodegenerative Diseases, vol. 6, no. 2, pp. 9–14.

Imbimbo, B. P., Solfrizzi, V., & Panza, F. 2010, “Are NSAIDs useful to treat Alzheimer’s disease or mild cognitive impairment?.” Frontiers in aging neuroscience, vol. 2, pp. 19.

Jamsen, K.M.P.D., Ilomaki, J.P.D., Hilmer, S.N.P.D., Jokanovic, N.B.P., Tan, E.C.K.P.D. & Bell, S.J.P.D. 2016, “A systematic review of the statistical methods in prospective cohort studies investigating the effect of medications on cognition in older people”, Research In Social & Administrative Pharmacy, vol. 12, no. 1, pp. 20–28.

Joshi, A., Wang, D. H., Watterson, S., McClean, P. L., Behera, C. K., Sharp, T., & Wong-Lin, K. (2020). Opportunities for multiscale computational modelling of serotonergic drug effects in Alzheimer’s disease. Neuropharmacology, 174, 108118.

Kaur, D., Bucholc, M., Finn, D.P., Todd, S., Wong-Lin, K. & McClean, P.L. 2020, “Multi-time-point data preparation robustly reveals MCI and dementia risk factors.”, Alzheimer’s & Dementia : Diagnosis, Assessment & Disease Monitoring, vol. 12, no. 1, pp. e12116.

Kim, J.Y., Ku, Y.S., Kim, H.J., Trinh, N.T., Kim, W., Jeong, B., Heo, T.Y., Lee, M.K. & Lee, K.E. 2019, “Oral diabetes medication and risk of dementia in elderly patients with type 2 diabetes.”, Diabetes Research & Clinical Practice, vol. 154, pp. 116–123.

Kotilinek, L. A., Westerman, M. A., Wang, Q., Panizzon, K., Lim, G. P., Simonyi, A., Lesne, S., Falinska, A., Younkin, L. H., Younkin, S. G., Rowan, M., Cleary, J., Wallis, R. A., Sun, G. Y., Cole, G., Frautschy, S., Anwyl, R., & Ashe, K. H. 2008. Cyclooxygenase-2 inhibition improves amyloid-beta-mediated suppression of memory and synaptic plasticity. Brain : a journal of neurology, 131(Pt 3), 651–664.

Kukull, W.A. (2015) NACC UNIFORM DATA SET. Researchers Data Dictionary. Available at: https://files.alz.washington.edu/documentation/uds3-rdd.pdf (Accessed: 05.03.2022).

Lee, Z. X., Ang, E., Lim, X. T., & Arain, S. J. 2021. Association of Risk of Dementia With Direct Oral Anticoagulants Versus Warfarin Use in Patients With Non-valvular Atrial Fibrillation: A Systematic Review and Meta-analysis. Journal of cardiovascular pharmacology, 77(1), 22–31.

Livingston, G., Huntley, J., Sommerlad, A., Ames, D., Ballard, C., Banerjee, S., Brayne, C., Burns, A., Cohen-Mansfield, J., Cooper, C., Costafreda, S.G., Dias, A., Fox, N., Gitlin, L.N., Howard, R., Kales, H.C., Kivimaki, M., Larson, E.B., Ogunniyi, A., Orgeta, V., Ritchie, K., Rockwood, K., Sampson, E.L., Samus, Q., Schneider, L.S., Selbaek, G., Teri, L. & Mukadam, N. 2020, “Dementia prevention, intervention, and care: 2020 report of the Lancet Commission”, Lancet, vol. 396, no. 10248, pp. 413–446.

McCombe, N., Liu, S., Ding, X., Prasad, G., Bucholc, M., Finn, D. P., Todd, S., McClean, P. L., & Wong-Lin, K. (2022). Practical Strategies for Extreme Missing Data Imputation in Dementia Diagnosis. IEEE journal of biomedical and health informatics, 26(2), 818–827. https://doi.org/10.1109/JBHI.2021.3098511

McKhann, G., Drachman, D., Folstein, M., Katzman, R., Price, D. & Stadlan, E.M. 1984, “Clinical diagnosis of Alzheimer’s disease: report of the NINCDS-ADRDA Work Group under the auspices of Department of Health and Human Services Task Force on Alzheimer’s Disease.”, Neurology, vol. 34, no. 7, pp. 939–944.

McKhann, G.M., Knopman, D.S., Chertkow, H., Hyman, B.T., Jack, C.R.J., Kawas, C.H., Klunk, W.E., Koroshetz, W.J., Manly, J.J., Mayeux, R., Mohs, R.C., Morris, J.C., Rossor, M.N., Scheltens, P., Carrillo, M.C., Thies, B., Weintraub, S. & Phelps, C.H. 2011, “The diagnosis of dementia due to Alzheimer’s disease: recommendations from the National Institute on Aging-Alzheimer’s Association workgroups on diagnostic guidelines for Alzheimer’s disease.”, Alzheimer’s & Dementia, vol. 7, no. 3, pp. 263–269.

Moore, E.M., Mander, A.G., Ames, D., Kotowicz, M.A., Carne, R.P., Brodaty, H., Woodward, M., Boundy, K., Ellis, K.A., Bush, A.I., Faux, N.G., Martins, R., Szoeke, C., Rowe, C., Watters, D.A. & AIBL Investigators 2013, “Increased risk of cognitive impairment in patients with diabetes is associated with metformin.”, Diabetes care, vol. 36, no. 10, pp. 2981–2987.

Morris, J.C. 1993, “The Clinical Dementia Rating (CDR): Current version and scoring rules”, Neurology, vol. 43, no. 11, pp. 2412–2414.

Morris, J.C., Weintraub, S., Chui, H.C., Cummings, J., Decarli, C., Ferris, S., Foster, N.L., Galasko, D., Graff-Radford, N., Peskind, E.R., Beekly, D., Ramos, E.M. & Kukull, W.A. 2006, “The Uniform Data Set (UDS): clinical and cognitive variables and descriptive data from Alzheimer Disease Centers.”, Alzheimer Disease & Associated Disorders, vol. 20, no. 4, pp. 210–216.

Newcombe, E.A., Camats-Perna, J., Silva, M.L., Valmas, N., Huat, T.J. & Medeiros, R. 2018, “Inflammation: the link between comorbidities, genetics, and Alzheimer’s disease”, Journal of Neuroinflammation, vol. 15, no. 1, pp. 276.

Nikaido A.M., Ellinwood Jr. E.H., Heatherly D.G. & Gupta, S.K. 1990, “Age-related increase in CNS sensitivity to benzodiazepines as assessed by task difficulty.”, Psychopharmacology, vol. 100, no. 1, pp. 90–97.

Nishtala P.S., Salahudeen M.S. & Hilmer, S.N. 2016, “Anticholinergics: theoretical and clinical overview.”, Expert Opinion on Drug Safety, vol. 15, no. 6, pp. 753–768.

Obermann K.R., Morris J.C. & Roe, C.M. 2013, “Exploration of 100 commonly used drugs and supplements on cognition in older adults.”, Alzheimer’s and Dementia, vol. 9, no. 6, pp. 724–732.

O’Bryant S.E., Lacritz L.H., Hall J., Waring S.C., Chan W., Khodr Z.G., Massman P.J., Hobson V. & Cullum, C.M. 2010, “Validation of the new interpretive guidelines for the clinical dementia rating scale sum of boxes score in the National Alzheimer’s Coordinating Center database.”, Archives of Neurology, vol. 67, no. 6, pp. 746–749.

Ou, Y., Tan, C., Shen, X., Xu, W., Hou, X., Dong, Q., Tan, L. & Yu, J. 2020, “Blood Pressure and Risks of Cognitive Impairment and Dementia: A Systematic Review and Meta-Analysis of 209 Prospective Studies”, Hypertension, vol. 76, no. 1, pp. 217–225.

Peters, R., Booth, A., Rockwood, K., Peters, J., D’Este, C. & Anstey, K.J. 2019, “Combining modifiable risk factors and risk of dementia: a systematic review and meta-analysis.”, BMJ Open, vol. 9, no. 1, pp. e022846.

Petersen R.C. & Morris, J.C. 2005, “Mild cognitive impairment as a clinical entity and treatment target.”, Archives of Neurology, vol. 62, no. 7, pp. 1160–1163.

Raj, D., Yin, Z., Breur, M., Doorduin, J., Holtman, I.R., Olah, M., Mantingh-Otter, I.J., Van Dam, D., De Deyn, P.P., den Dunnen, W., Eggen, B.J.L., Amor, S. & Boddeke, E. 2017, “Increased White Matter Inflammation in Aging-and Alzheimer’s Disease Brain.”, Frontiers in Molecular Neuroscience, vol. 10, pp. 206.

Rouch, L., Cestac, P., Hanon, O., Cool, C., Helmer, C., Bouhanick, B., Chamontin, B., Dartigues, J., Vellas, B. & Andrieu, S. 2015, “Antihypertensive drugs, prevention of cognitive decline and dementia: a systematic review of observational studies, randomized controlled trials and meta-analyses, with discussion of potential mechanisms.”, CNS Drugs, vol. 29, no. 2, pp. 113–130.

Salahudeen M.S., Duffull S.B. & Nishtala, P.S. 2015, “Anticholinergic burden quantified by anticholinergic risk scales and adverse outcomes in older people: a systematic review.”, BMC geriatrics, vol. 15, pp. 31.

Salzman, C. 2020, “Do Benzodiazepines Cause Alzheimer’s Disease?.”, American Journal of Psychiatry, vol. 177, no. 6, pp. 476–478.

Schneider, L.S., Insel, P.S., Weiner, M.W. & for the Alzheimer’s Disease Neuroimaging Initiative 2011, “Treatment With Cholinesterase Inhibitors and Memantine of Patients in the Alzheimer’s Disease Neuroimaging Initiative”, Archives of Neurology, vol. 68, no. 1, pp. 58–66.

Schultz, B. G., Patten, D. K., & Berlau, D. J. 2018. The role of statins in both cognitive impairment and protection against dementia: a tale of two mechanisms. Translational neurodegeneration, 7, 5.

Seaquist, E. R., Miller, M. E., Fonseca, V., Ismail-Beigi, F., Launer, L. J., Punthakee, Z., & Sood, A. 2013. Effect of thiazolidinediones and insulin on cognitive outcomes in ACCORD-MIND. Journal of diabetes and its complications, 27(5), 485–491.

Shah, K., Qureshi, S.U., Johnson, M., Parikh, N., Schulz, P.E. & Kunik, M.E. 2009, “Does use of antihypertensive drugs affect the incidence or progression of dementia? A systematic review.”, American Journal of Geriatric Pharmacotherapy, vol. 7, no. 5, pp. 250–261.

Trenaman, S.C., Rideout, M. & Andrew, M.K. 2019, “Sex and gender differences in polypharmacy in persons with dementia: A scoping review.”, SAGE Open Medicine, vol. 7, pp. 2050312119845715.

Verdoux, H., Lagnaoui, R. & Begaud, B. 2005, “Is benzodiazepine use a risk factor for cognitive decline and dementia? A literature review of epidemiological studies.”, Psychological medicine, vol. 35, no. 3, pp. 307–315.

Wancata, J., Borjesson-Hanson, A., Ostling, S., Sjogren, K. & Skoog, I. 2007, “Diagnostic criteria influence dementia prevalence.”, American Journal of Geriatric Psychiatry, vol. 15, no. 12, pp. 1034–1045.

Woodling, N. S., Colas, D., Wang, Q., Minhas, P., Panchal, M., Liang, X., Mhatre, S. D., Brown, H., Ko, N., Zagol-Ikapitte, I., van der Hart, M., Khroyan, T. V., Chuluun, B., Priyam, P. G., Milne, G. L., Rassoulpour, A., Boutaud, O., Manning-Boğ, A. B., Heller, H. C., & Andreasson, K. I. 2016. Cyclooxygenase inhibition targets neurons to prevent early behavioural decline in Alzheimer’s disease model mice. Brain : a journal of neurology, 139(Pt 7), 2063–2081. https://doi.org/10.1093/brain/aww117

Wu C.Y., Ouk M., Wong Y.Y., Anita N.Z., Edwards J.D., Yang P., Shah B.R., Herrmann N., Lanctot K.L., Kapral M.K., MacIntosh B.J., Rabin J.S., Black S.E. & Swardfager, W. 2020, “Relationships between memory decline and the use of metformin or DPP4 inhibitors in people with type 2 diabetes with normal cognition or Alzheimer’s disease, and the role APOE carrier status.”, Alzheimer’s and Dementia, vol. 16, no. 12, pp. 1663–1673.

Xiong, G., Yang, Z., Yi, J., Wang, N., Wang, L., Zhu, H., Wu, C., Lu, A., Chen, X., Liu, S., Hou, T., & Cao, D. 2022. DDInter: an online drug-drug interaction database towards improving clinical decision-making and patient safety. Nucleic acids research, 50(D1), D1200–D1207.

Zammitt, N.N.M. & Frier, B.M. 2005, “Hypoglycemia in Type 2 Diabetes: Pathophysiology, frequency, and effects of different treatment modalities”, Diabetes care, vol. 28, no. 12, pp. 2948–2961.

Zeng, D., Jiang, C., Su, C., Tan, Y., & Wu, J. (2019). Anticoagulation in atrial fibrillation and cognitive decline: A systematic review and meta-analysis. Medicine, 98(7), e14499.

Zhang, C., Wang, Y., Wang, D., Zhang, J. & Zhang, F. 2018, “NSAID Exposure and Risk of Alzheimer’s Disease: An Updated Meta-Analysis From Cohort Studies.”, Frontiers in aging neuroscience, vol. 10, pp. 83.

